# ROLE OF LUNG ULTRASOUND FOR THE ETIOLOGICAL DIAGNOSIS OF COMMUNITY- ACQUIRED PNEUMONIA IN CHILDREN: A PROSPECTIVE STUDY

**DOI:** 10.1101/2020.10.31.20223867

**Authors:** Danilo Buonsenso, Annamaria Musolino, Valentina Ferro, Cristina De Rose, Rosa Morello, Chiara Ventola, Flora Marzia Liotti, Antonio Chiaretti, Daniele Guerino Biasucci, Teresa Spanu, Maurizio Sanguinetti, Piero Valentini

**Affiliations:** Department of Woman and Child Health and Public Health, Fondazione Policlinico Universitario A. Gemelli, Rome, Italy; Dipartimento di Scienze Biotecnologiche di Base, Cliniche Intensivologiche e Perioperatorie, Università Cattolica del Sacro Cuore, Rome, Italy; Global Health Research Institute, Istituto di Igiene, Università Cattolica del Sacro Cuore, Roma, Italia; Department of Pediatric Emergency Medicine, Bambino Gesù Children’s Hospital, IRCCS, Rome, Italy; Department of Anesthesia and Intensive Care, Fondazione Policlinico Universitario “A.Gemelli,” Rome, Italy; Dipartimento di Scienze di Laboratorio e Infettivologiche, Fondazione Policlinico Universitario A. Gemelli IRCCS, Rome, Italy

**Keywords:** lung ultrasound, pneumonia, CAP, etiological diagnosis, children

## Abstract

**Objective and design:** Our prospective study is one of the largest prospective study assessing the role of detailed lung ultrasound features to discriminate the etiological diagnosis of Community acquired pneumonia (CAP) in children.

**Methodology:** We prospectively analysed patients aged from 1 month to 17 years admitted between March 2018 and April 2020 who were hospitalized for CAP. For all patients included in the study, history, clinical parameters, microbiological data, and lung ultrasound data were collected.

Patients were stratified into three main groups (“bacterial”, “viral”, “atypical”) according to the presumed microbial aetiology and lung ultrasound findings evaluated according to the aetiological group.

**Results:** We found that some ultrasound findings as size, number and distribution of consolidations, the position and motion of air bronchograms, pleural effusions and distribution of vertical artifacts significantly differ (p < 0.05) in children with bacterial, viral and atypical CAP. Conversely, clinical parameters and laboratory were not able to significantly distinguish between these groups. Chest x-ray, despite being still widely used, was the less useful tool in this discrimination.

**Conclusion:** Our study provides a detailed analysis of LUS features able to predict the etiology CAP in children. These findings may help the physicians to better manage a child with CAP and to offer personalized approach, from diagnosis to treatment and follow-up.

## INTRODUCTION

Community acquired pneumonia (CAP) represents the single largest cause of paediatric morbidity and mortality worldwide.^1,2^ The diagnosis of pneumonia is essentially based on medical history and clinical examination, which has poor sensitivity and specificity^3,4^ and do not allow to identify the causative agent responsible for the infection.^5^

Although viruses represent the most frequent cause of CAP, the majority of children with suspected or confirmed CAP are still treated with empirical and often unnecessary antibiotics^6^, contributing to the spread of antibiotic resistance, one of the biggest medical emergencies of modern medicine. In the Centers for Disease Control and Prevention Etiology of Pneumonia in the Community cohort, only 15% of hospitalized children with radiographic pneumonia had a detectable bacterial aetiology; however, 88% received antibiotics.^7^ A recently published among Pakistani children younger than 5 years of age with pneumonia with tachypnea randomized at amoxicillin vs placebo, the number of children with pneumonia and tachypnea who would have needed to be treated with amoxicillin to prevent one treatment failure was 44, suggesting that a significant number of CAP is of viral origin and does not requires antibiotic.^8^

However, current guidelines do not help the physician on how to approach to an optimised strategy for prevention, diagnosis and treatment of disease for each single child with CAP, based on his or her unique characteristics, but mainly suggest a general approach to paediatric CAP.^9^

To date, both clinical findings^10^ and laboratory results^10-16^ failed to accurately distinguish viral, bacterial and atypical pneumonia. Even studies that report differences in laboratory biomarkers could not determine any reliable thresholds for differentiating bacterial pneumonia from viral pneumonia^13^, since normal tests do not always exclude bacterial CAP.^9^

Chest x-ray (CXR) is not necessary to confirm the diagnosis of CAP in milder cases, who are treated as outpatients and is also associated with risk of radiation exposure.^17^ Moreover, CXR cannot reliably establish the microbial diagnosis of CAP^18,19-22^ and the interpretation of radiographic images varies significantly among the observers.^23^

In recent years, lung ultrasound (LUS) use has been widely studied as an alternative diagnostic tool for CAP of both bacterial and viral origin, proving to have high specificity and sensitivity for the diagnosis and as regards the follow-up in children with pneumonia.^21,24-26^ Moreover, LUS has several advantages over X-ray, particularly useful for the paediatric population: radiation-free, lower cost, possibility of follow-up examinations, ability to monitor treatment, easy accessibility in all settings (including poor countries), fast, and can be used immediately as a point-of-care method. LUS results are immediately available to the clinician, who must decide about the initial empirical treatment.^19,21-26^

The new challenge of LUS is to determine the aetiology of CAP. First studies primarily aimed to assess the role of LUS in CAP suggested that smaller consolidations may be associated with a viral aetiology^27^, but only one study prospectively assessed the role of LUS in defining the aetiology of CAP.^19^ However, although this study found that small subpleural consolidations and/or an increased number of B-lines (interstitial syndrome) are characteristics of viral pneumonia, the authors did not assessed important LUS features (such as the bronchograms, type and location of vertical artifacts, type of effusions).

Therefore, due to the growing role of LUS in the evaluation of patients with respiratory conditions and the limits of available data, we carried out this prospective study aiming to assess the role of LUS in supporting the etiological diagnosis of CAP in children.

## PARTICIPANTS AND METHODS

### Study Population

We prospectively analysed patients aged from 1 month to 17 years admitted between March 2018 and April 2020 who were hospitalized for CAP and had pneumonic infiltrates detected with LUS. For all patients included in the study, history, clinical parameters, microbiological data, and ultrasound data were collected. The study was approved by the Ethics Committee of Fondazione Policlinic Universitario A. Gemelli IRCCS, Rome, Italy. Informed consent was obtained by all participants and/or their legal guardians (for children younger than 16 years). There is no identifying information or image in the article.

### Patients

The evaluating physician made the clinical diagnosis of CAP in accordance with the British Thoracic Society guidelines.^18^ At the first evaluation in the Emergency Department (ED), all children with suspected CAP underwent medical history and clinical evaluation. Further investigations were performed only when deemed necessary from the evaluating paediatricians (anteroposterior CXR; blood tests including white cell count (WCC), C-reactive protein (CRP) and procalcitonin (PCT). The physician on duty made decisions about the patient’s diagnosis and treatment according to his/her own practice and without knowledge of the LUS findings, but aware of other clinical/laboratory/imaging data if performed. In our institution, the local protocol for CAP antibiotic treatment follows the Pediatric Infectious Diseases Society and the Infectious Diseases Society of America.^9^

#### CAP definitions

We defined as CAP those patients requiring: acute respiratory signs and symptoms, fever > 37.5°C, clinical or radiological evidence of a new pulmonary infiltrates.

#### Inclusion criteria

Children with a clinical diagnosis of CAP (based on history, clinical examination, blood tests (if performed), and CXR (if performed) who underwent LUS within six hours from the first clinical evaluation and with available clinical information about the outcome (including the etiological suspicion) available.

#### Exclusion criteria

Patients with underlying diseases, including respiratory tract anomalies, immunodeficiency, cerebral palsy, neuromuscular diseases, congenital heart disease, and malignancy were excluded, as well as children who did not undergo LUS at 48 hours after treatment.

#### Etiological stratifications of patients

Patients were stratified into different groups according to the presumed microbial etiology: patients with bacterial pneumonia, patients with viral pneumonia and patients positive for atypical pneumonia, especially *Mycoplasma pneumoniae*. An expert in pediatric infectious diseases assessed the final database, blinded to the clinical discharge charts, and classified the aetiological diagnosis according to the following data.

Bacterial CAP was considered in patients documented bacterial infection (either culture- or PCR-based methods) in clinically significant samples (bronchoalveolar lavage, pleural drainage, blood cultures), and/or lobar pneumonia on CXR (if performed), and/or leukocytosis (> 15 x 109/L), and/or raised inflammatory markers (either CRP or PCT, according to clinical decision and local availability), even when viruses were detected in the nasopharyngeal swab. When no viruses or atypical bacteria were detected, the clinical opinion of the responsible pediatrician, based on clinical, laboratory and imaging studies, was considered enough for the stratification into the bacterial CAP group.

Patients with detected viral infection on nasopharyngeal were included into the viral CAP group only after the exclusion of bacterial superinfection, according to a comprehensive assessment of clinical, laboratory, radiology and microbiological finding. Children were included into the atypical CAP group when *Mycoplasma pneumoniae* or *Chlamydia pneumoniae* were detected on nasopharyngeal swabs and bacterial superinfection was excluded, and clinical data were compatible with a diagnosis of atypical pneumonia.

### Microbiological studies

Nasopharyngeal swabs were obtained and tested fresh from all patients. MicrobScan, a qualitative diagnostic test developed by Nurex S.r.l. (Sassari, Italy) was used for the detection and identification of bacterial and fungal DNA in respiratory samples. The system consists of an open robotic platform, which performs in succession extraction of DNA and preparation of the plate for the RT-qPCR, up to a maximum of 24 samples in an automated way. Microbial DNA was extracted and amplified from a sample volume of 100 μl. The target species panel includes *Bordetella pertussis, Bordetella parapertussis, Legionella pneumophila, Chlamydia pneumoniae, Mycoplasma pneumoniae* and the fungus *Pneumocystis jirovecii*. The Seegene Allplex™ Respiratory Panel 1, 2 and 3 assays were used for the molecular diagnosis of viral respiratory pathogens. The organisms detected by the three panels include influenza virus A (subtypes H1, pdm09 and H3), influenza virus B, respiratory syncytial virus types A and B, adenovirus, metapneumovirus, human enterovirus, parainfluenza virus 1, parainfluenza virus 2, parainfluenza virus 3, parainfluenza virus. human bocavirus, coronavirus OC43, coronavirus 229E coronavirus, NL63 and human rhinovirus. In addition, bacterial culture of blood and respiratory samples were performed, when indicated.

### Lung ultrasound

LUS was performed with the ultrasound machine Esaote Mylab 40. It was performed within 6 hours from the clinical diagnosis of CAP, unaware of microbiological, laboratory and CXR (when performed) results. Linear probe (12 – 6 MHz) was used in preschool children. In older children, we used a curved probe (8 – 5 MHz).

Images and clips were stored and archived. All LUS was made by the same physician that made the first one to reduce inter-operator differences.

The following LUS features were recorded (Figure 1):

**FIGURE 1.**
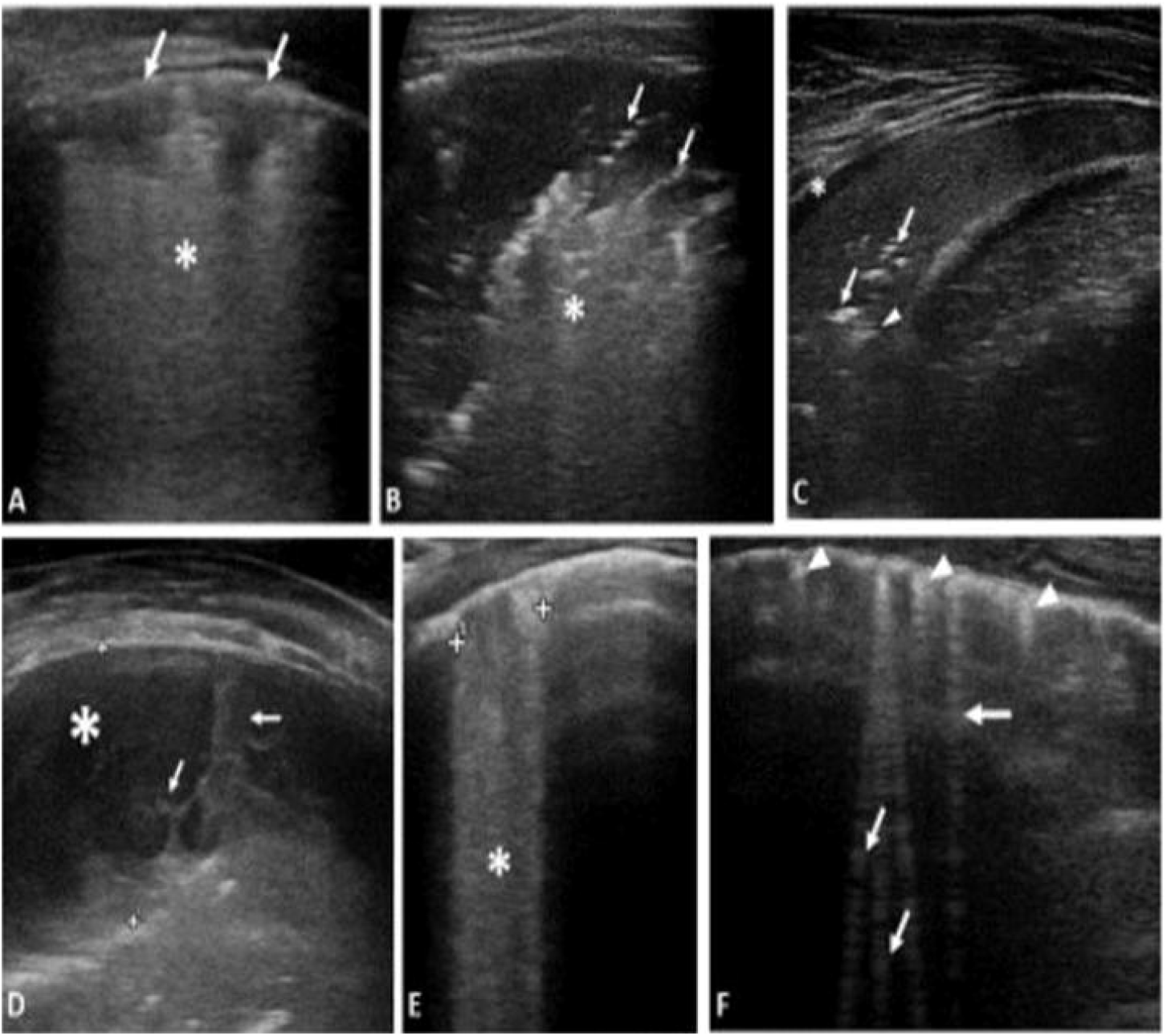
**A**. VIRAL PNEUMONIA (H1N1): Sub-centimeter subpleural consolidation *(arrows)* associated with vertical long perilesional vertical artifacts and areas of white lung *(asterisk)*. **B**. ATYPICAL PNEUMONIA (Mycoplasma pneumoniae): Subpleural consolidation of less than 4 cm in size with dynamic superficial air bronchogram *(arrow)* and perilesional and deep confluent vertical artifacts *(asterisk)*. **C**. BACTERIAL PNEUMONIA: Compact / hepatized large subpleural consolidation with static air bronchograms *(arrow)* and deep fluid bronchogram *(arrowhead)*. Minimum reactive pleural effusion *(asterisk)*. **D**. COMPLEX PLEURAL EFFUSION IN A BACTERIAL PNEUMONIA: complicated pleural effusion *(asterisk)* with multiple and concamerated fibrin *(arrow)*. **E**. Confluent long vertical artifacts *(asterisk)*. **F**. Isolated long vertical artifacts (*arrow*); short vertical artifacts (*arrowhead*).

– size of the main lesion, that we generally define as subpleural lung parenchymal lesion (Consolidation and Atelectasis), if they are single or multiple and location (monolateral or bilateral)
– presence of bronchograms, its characteristics (air or fluid), morphology (arboriform or dot□like/linear), position (deep if >2 cm far from the pleura or superficial if close to the pleura), dynamicity during breath (fix, poorly dynamic, or clearly dynamic);
– presence of vertical artifacts, charactheristics (short or long, spared or confluents), position (monolateral or bilateral, perilesional or not)
– presence and type of pleural effusion: simple (anechogenic and dependent to gravity) or complex (presence of septa, hyperechogenic spot, following the lung through the apex and not dependent to gravity, requiring drainage).

The scans were made by investigating the anterior, lateral and posterior regions of the thorax and placing the probe transversally and longitudinally along the lines considered traditional ultrasound findings: the parasternal line, the axillary line and the paravertebral line so as to fully explore the chest wall according to a methodical scheme first described by Copetti et al.^24^ To investigate the anterior and lateral lung fields patients were positioned, according to age, in a seated or supine position. The posterior lung fields have been explored in lateral decubitus and in sitting position. We compared the ultrasound results with those of the nasopharyngeal swab to verify the concordance of the etiological diagnosis, according to the three main groups “bacterial”, “viral”, “atypical”.

### Treatment

We considered standard of care (SOC) the first line treatments (for either outpatient or inpatient) as per international guidelines^9^ (amoxicillin, amoxicillin/clavulanate, ceftriaxone, cefotaxime, ampicillin, and penicillin).

### Statistical analysis

In order to determine the power of the study, the study was set up as a study of agreement between lung ultrasound and the aetiology of CAP. For atypical pneumonia there is a 60% agreement (then a 40% disagreement); for bacterial pneumonia, the agreement rises to 90% (disagreement 10%). To be conservative we considered the highest percentage of disagreement that is 40%. In this case considering alpha= 0.01 and beta= 0.20, N= 159. A Statistical analysis was performed using the software STATA/IC 14.2 version 2017. We use the Skewness/Kurtosis test to verify the normality of the distribution. Continuous variables showed non-parametric distribution and were presented as median [interquartile range (IQR)]. Categorical variables were reported as frequency and percentage. Categorical variables were compared using chi-square or Fisher’s exact test, as appropriate. Continuous variables were assessed using Mann–Whitney U. We performed the Multivariable logistic regression analysis to identify potential predictors, among US features, of bacterial infection compared with viral infection, viral infection compared with atypical infection and bacterial infection compared with atypical, adjusted for age, sex, clinical and laboratory characteristics. We considered a 2-tailed p value less than 0.05 to be significant. The dataset is available upon reasonable request. The interrater reliability for each variable was with the Cohen’s k coefficient. The concordance was considered absent for k values lower than 0, poor if between 0 and 0.4; discrete if between 0.4 and 0.6, good if k between 0.6 and 0.8; high if above 0.8

## RESULTS

### Patients’ characteristics

A total of 186 children with suspected CAP (103 male and 83 female) with an average age of six were enrolled in the study (Figure 2). The prevalent etiology of CAP was viral, with a slight prevalence in male (55% of the cases). Main epidemiological and clinical characteristics of the study population, assessed according to the aetiological group, are described in detail in Table 1. As reported, the majority of clinical parameters were not able to discriminate the different aetiological groups.

**FIGURE 2.**
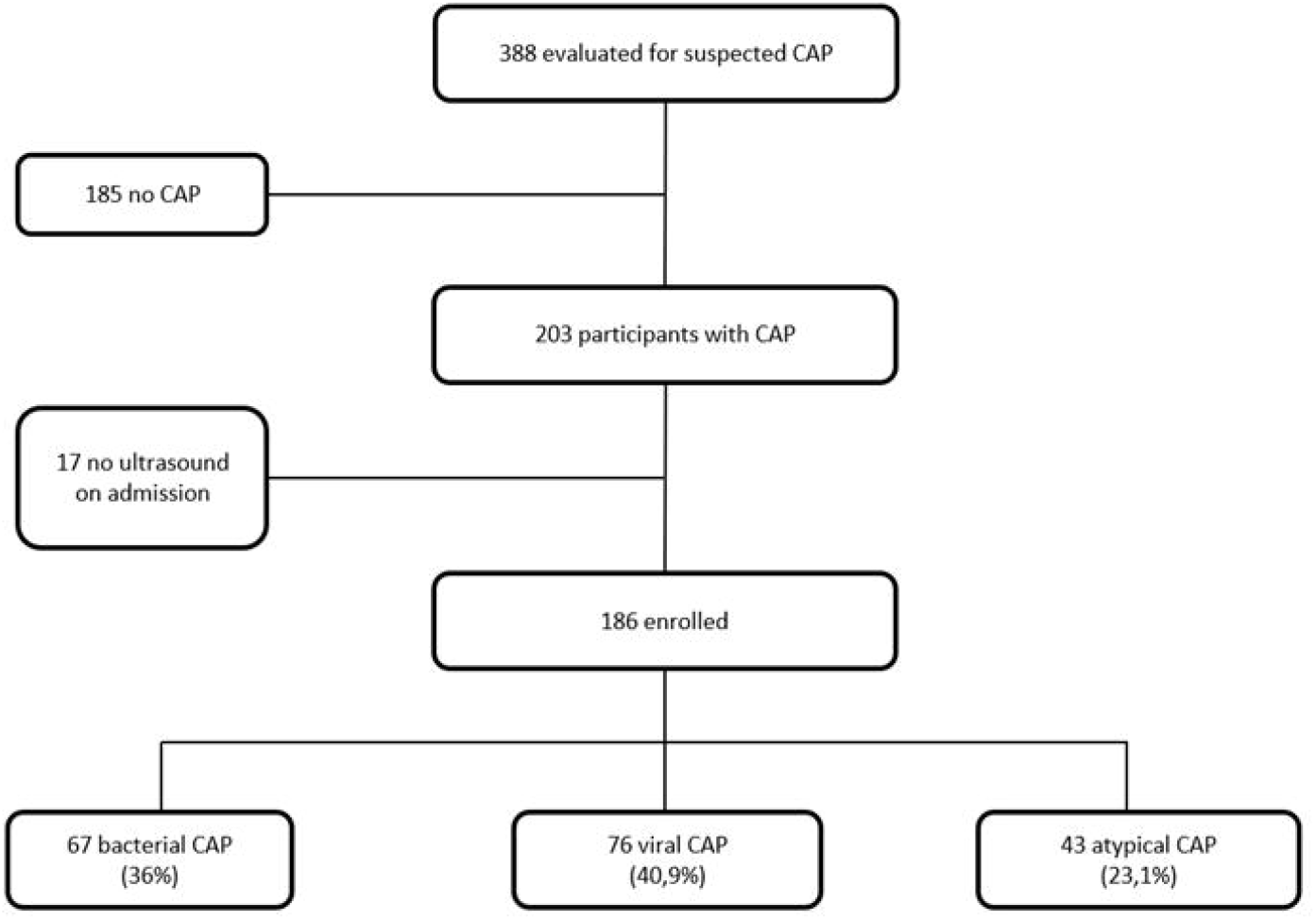
Study flow.

Main diagnostic and laboratory investigations, assessed according to the aetiological group, are described in detail in Table 2. Neither laboratory nor CXR findings were able to significantly discriminate the etiological groups since, also in case of statistical significance, the overlap between etiological groups was evident.

### LUS findings

Details about LUS features in children with bacterial, viral and atypical pneumonia are described in table 3. Consolidations were found in 100% of bacterial infections, with a significant statistical difference compared to viral infections (P value= 0.001) and to atypical infections (P value=0.004). We analyzed the size of consolidation according to the etiological group:

– for consolidation <1.5 cm, there is a statistical significance between bacterial and viral infections (P value= <0.001) and between bacterial and atypical infections (P value= 0.004). They were more expressed in viral pneumonia (63.8%);
– for consolidation between 1.5 and 4 cm, there is a statistical significance between bacterial and viral infections (P 0.022), and they were more expressed in bacterial pneumonia (55.22%);
– for consolidation >4cm, there is a statistical significance between bacterial and viral infections (P value <0.001) and between bacterial and atypical infections (P value <0.001). They were more expressed in bacterial infections (22.39%) and poorly expressed in viral infections (1.54%).

Multiple and bilateral consolidation are infrequent in bacterial infections, making them statistically significant to distinguish bacterial pneumonia from viral and atypical diseases (P < 0.0001). We also considered air bronchogram, that was highly represented in bacterial (77.61%) and atypical (60.47%) CAP. There is a statistical significant difference between bacterial infections and viral infections (P value= <0.001) and between viral infections and atypical infections (P value= 0.004). Different characteristics of the air bronchogram have been taken into consideration:

– position: deep air bronchogram was found in 53.85% of bacterial pneumonia and 56% of viral pneumonia; superficial air bronchogram was almost always present in the atypical pneumonia (76.92%). Position is a valid statistical parameter to distinguish atypical infection from the bacterial one (P value=0.010) and the viral one (P value= 0.010); however, it is not a statistically significant parameter for distinguish between the bacterial and viral pneumonia (P value= 0.859);
– dynamic bronchogram was often found in atypical infections (69.23%) and moderately represented in bacterial (44%) and viral (37.50%) infections. Static bronchogram were found in 56% of cases in bacterial forms, 62.50% of cases in viral forms and in 30.77% of cases in atypical forms (P > 0.05).
– the presence of fluid bronchogram was more frequently described in case of bacterial pneumonia compared with viral (P <0.001) and atypical pneumonia (P 0.003).

Pleural effusions were significantly more frequent in children with bacterial CAP (P < 0.0001). Complicated effusion was not represented in viral forms.

We also assessed vertical deep artefacts. They were frequently found in the viral (88.16%) and atypical (88.37%) infections, and moderately found in the bacterial ones (58.21%). Children with viral and atypical pneumonia had significantly more vertical deep artefacts compared with bacterial pneumonia (P <0.001 and P 0.001, respectively), while there were no significant differences between viral and atypical pneumonia (P > 0.05). Also the distribution of the vertical deep artefacts in the three groups, being significantly more diffuse in children with viral (78.57%) and atypical (70.27%) CAP (P < 0.05), while children with bacterial CAP mainly had vertical artefacts located in continuity with the main consolidation.

Tables 4, 5 and 6 shows the multivariable logistic regression analyses applied to compared the three main groups.

#### Interobserver agreement

We made interobserver agreement on a random set of 10% images. The levels of interobserver agreement were good (values of coefficient between 0.6 and 0.8) for air bronchogram position (deep, superficial) and motion (static, dynamic), while was above 0.8 (confirming a high interobserver agreement) for consolidation size, effusions, presence and distribution of vertical artifacts.

## DISCUSSION

Our prospective study is one of the largest prospective study assessing the role of detailed LUS features to discriminate bacterial, viral and atypical CAP in children. We found that the size, number and distribution of consolidations, the position and motion of air bronchograms, pleural effusions and distribution of vertical artifacts significantly differ in children with bacterial, viral and atypical CAP. Such differences were particularly relevant when bacterial CAPs were compared with viral and atypical ones. Conversely, clinical parameters, including fever, chest pain and main auscultation features, and laboratory were not able to significantly distinguish between these groups. CXR, despite being still widely used, was the less useful tool in this discrimination. These findings highlight, therefore, the need of new tools to support the etiological classification of pediatric CAP and we proved that several LUS features may easily support clinicians in this.

Given the increasing phenomenon of antibiotic resistance in children, the possibility of knowing the etiology of the infection represents an important step forward. The first decade of LUS studies focused on the role of LUS in detecting pneumonia. A recently performed meta-analysis confirmed high sensitivity (96%) and specificity (93%) of LUS for detecting pneumonia in children.^4^ First studies showed specific LUS patterns to diagnose viral lower respiratory tract infections and bronchiolitis in children.^28-31^. Buonsenso and colleagues^15,24^ showed that specific LUS patterns on diagnosis and after 48 hours of treatments (bronchograms, consolidation size, characteristics of pleural effusion) were predictive of antibiotic response in children with CAP, more than clinical data and laboratory results. Moreover, recent basic science and clinical studies are clarifying the knowledge of and genesis of artifacts generated by the ultrasound-lung interaction. In particular, performing physical analyses on artificial models and using modern deep learning strategies and train a fully convolutional neural network, Demi and colleagues^32,33,34^ showed that B-lines have different morphologies according to the medical conditions that generate them (interstitial lung diseases, cardiogenic and non-cardiogenic lung edema, interstitial pneumonia and lung contusion). These data, all together suggested that a more detailed analyses of LUS features (“ultrasonomic”) may represent a new and promising tool to personalize the diagnosis, treatment, follow-up and care of children with CAP. In this context, recognizing bacterial pneumonia from a viral or atypical using LUS at patient’s bed-side would allow us to offer a more focused approach and treatment to each child, responding to a modern medical concept that each patient is a unique one and requires a personalized approach. To date, few studies have described LUS findings able to define the etiology of pediatric. In particular, Berce et al^19^ evaluated 147 children hospitalized because of CAP, showing that LUS detected consolidations in viral CAP were significantly smaller, with a median diameter of 15mm, compared to 20mm in atypical bacterial LRTIs (p = 0.05) and 30 mm in bacterial LRTIs (p <0.001). Other authors also highlighted that consolidation size or distribution can support the diagnosis of viral bronchiolitis, Influenza pneumonia and COVID19 pneumonia.^21,28-30,35^ These findings were comparable with our study. However, our study has evidenced more important LUS features not previously assessed. We found that air bronchograms were more common in bacteria and atypical CAP but, importantly, fluid bronchograms were almost exclusively described in bacterial cases. Also, complicated pleural effusions were never described in viral CAPs. Vertical artefacts, which gained more interest during the last year and in particular since LUS has been routinely used in COVID-19 pneumonia^21,35-38^, also played a significant role, since in bacterial CAP were mainly located in proximity of the main consolidation, while in the others were mostly diffuse and bilateral.

Considering the well-known advantages of LUS (a low-cost, easily reproducible, non-invasive tool that does not cause pain and damage induced by radiations), all the ultrasound findings obtained by our study can have an important impact on the daily pediatric clinical practice. LUS, particularly when used in adjunction to clinical examination by experienced operators, can support the etiological of pneumonia and offer a personalized approach to patients. If further confirmed, this approach can also support antibiotic stewardship programs.

Importantly, the detailed analyses offered by our study represent a modern view of LUS, as recently highlighted by international experts^39^: during the last decades, LUS moved from being a diagnostic tool (qualitative approach) to a monitoring tool for lung aeration quantification (i.e.,quantitative approach). In this new vision, a more attention to several LUS features (vertical artifacts, consolidations, echogenicity, bronchograms, and dynamic data) need to be taken in consideration. Artificial intelligence softwares, since we are entering the era of the digital medicine (a process particularly speed-up by the COVID-19 pandemic), will also support this view.^40^

Our study has limitations to address. The gold standard for the diagnosis of CAP is the chest CT scan, however its routine use in children is not ethical in children so we did not do the CT. Therefore, the diagnosis of CAP for the inclusion of patients in our study was made on a clinical bases according to the available guidelines. The stratification of patients according to the etiology cannot be 100% accurate, since the definitive microbiological diagnosis in non-mechanically ventilated children is difficult to establish, since bronchoalveolar lavages are invasive procedures. However, to date all studies assessing the microbiological diagnoses of CAP, including vaccine-probe study^41^, have the same limitation that is difficult to address. Nevertheless, we used a comprehensive assessment of each child including clinical, laboratory, imaging and microbiological data, including last generation molecular assays. On this regard, a limit is that we did not use quantitative assessments of nasopharyngeal PCR results that, according to recent studies in adult patients, have been positively associated with a diagnosis of CAP other than simple colonization.^42^

In conclusion, our study provides a detailed analysis of LUS features able to predict the etiology CAP in children. These findings may help the physicians to better manage a child with CAP and to offer personalized approach, from diagnosis to treatment and follow-up. However, further studies on pediatric CAP are necessary to confirm our findings.

## Data Availability

All data referred to in the manuscript and note links below.

