## Supplementary material for "ROLE OF LUNG ULTRASOUND FOR THE ETIOLOGICAL DIAGNOSIS OF COMMUNITY- ACQUIRED PNEUMONIA IN CHILDREN: A PROSPECTIVE STUDY": Tables

| Epidemiological/clinical  characteristics | Bacterial Infection  n=67 | Viral Infection  n=76 | Atypical Infection  n=43 | *P value* |
| --- | --- | --- | --- | --- |
| Gender  Female  Male | 27 (40.30)  40 (59.70) | 34 (44.74)  42 (55.26) | 22 (51.16)  21 (48.84) | B-V: 0.592  V-A: 0.5  B-A:0.263 |
| Age (years, median, SD) | 4 (7) | 2 (5.5) | 7 (5) | **B-V: 0.011**  **V-A:<0.001**  **B-A: 0.002** |
| Fever | 56 (83.54) | 61 (80.26) | 32 (74.42) | B-V: 0.608  V-A:0.459  B-A: 0.241 |
| Cough | 40 (59.70) | 51 (67.11) | 37 (86.05) | B-V: 0.358  **V-A: 0.024**  **B-A:0.003** |
| Chest pain | 10 (14.93) | 4 (5.26) | 4 (9.30) | B-V: 0.05  V-A:0.398  B-A:0.282 |
| Respiratory distress | 19 (28.79) | 20 (26.67) | 3 (6.98) | B-V: 0.779  **V-A:0.007**  **B-A:0.004** |
| Wheezing  (at auscultation) | 16 (25) | 27 (36) | 11 (25.54) | B-V:0.162  V-A:0.224  B-A:0.94 |
| Crackles  (at auscultation) | 26 (40.63) | 21 (28.38) | 20 (46.51) | B-V: 0.130  V-A:0.047  B-A:0.546 |
| Decreased air entry  (at auscultation) | 46 (71.88) | 34 (45.95) | 23 (53.49) | **B-V: 0.002**  V-A:0.431  B-A:0.05 |
| Need Oxygen (low flow) | 19 (28.36) | 21 (27.63) | 3 (6.98) | B-V: 0.9  **V-A:0.005**  **B-A: 0.005** |
| Need Oxygen (HNFC) | 12 (17.21) | 18 (23.68) | 8 (18.60) | B-V: 0.4  V-A: 0.519  B-A:0.927 |
| Need Oxygen (CPAP) | 4 (5.97) | 4(5.96) | 1 (2.33) | B-V: 0.85  V-A:0.403  B-A:0.348 |
| Need Intubation | 8 (11.94) | 2 (2.63) | 0 | **B-V: 0.031**  V-A:0.406  **B-A: 0.016** |
| Need Intensive Care | 4(6.06) | 1 (1.32) | 0 | B-V:0.142  V-A:0.639  B-A: 0.130 |
| Complicated clinical course | 26 (38.81) | 6 (7.89) | 1 (2.33) | **B-V: <0.001**  V-A:0.207  **B-A:<0.001** |
| Medical disposition  Hospitalization | 64 (95.52) | 65 (85.53) | 32 (74.44) | **B-V: 0.045**  V-A:0.134  **B-A:0.001** |
| Length of hospitalization (days) | 8 (12) | 5 (4) | 3 (5) | **B-V:<0.001**  **V-A: 0.0025**  **B-A: < 0.001** |
| Length of therapy (days) | 10 (10) | 7 (5) | 7 (5) | **B-V<0.001**  **V-B: 0.07**  **B-A: <0.001** |

**Table 1.** Differences of the epidemiological and clinical features in community acquired pneumonia with three etiological causative agents.

| Diagnostic /laboratory  investigations | Bacterial Infection  n=67 | Viral Infection  n=76 | Atypical Infection  n=43 | *P value* |
| --- | --- | --- | --- | --- |
| Performed chest X-ray (CXR) | 64(95.52) | 63 (84) | 33 (76.74) | B-V: 0.026  V-A:0.330  B-A:0.003 |
| Consolidation (CXR) | 58 (90.63) | 49 (77.78) | 27 (81.82) | B-V: 0.047  V-A: 0.643  B-A:0.212 |
| Atelectasis (CXR) | 12 (18.75) | 12 (19.05) | 3(9.09) | B-V: 0.9  V-A:0.164  B-A:0.172 |
| Pleural effusion (CXR) | 39 (60.94) | 15 (23.81) | 9 (27.27) | B-V: **<0.001**  V-A:0.710  **B-A:0.002** |
| CRP <50 mg/L | 24 (40.68) | 52 (85.25) | 15 (65.22) | **B-V: <0.001**  V-B: 0.042  B-A: 0.046 |
| CRP 50-100 mg/L | 12 (20.34) | 3 (4.92) | 3 (13.04) | **B-V:0.01**  V-B: 0.202  B-A:0.336 |
| CRP > 100 mg/L | 23 (38.98) | 6 (9.84) | 5 (21.74) | **B-V:<0.001**  V-B:0.141  B-A:0.139 |
| WBC count:  <10,000/ µL | 23 (41.07) | 23 (42.59) | 9 (37.50) | B-V:0.872  V-B: 0.673  B-A:0.766 |
| WBC count:  10,000-15,000/ µL | 15 (26.79) | 18 (33.33) | 8 (33.33) | B-V:0.454  V-B:1  B-A:0.553 |
| WBC count:  >15,000/ µL | 18 (32.14) | 13(24.07) | 7 (29.17) | B-V: 0.347  V-A:0.635  B-A:0.792 |

**Table 2.** Diagnostic/laboratory investigations

| Characteristics lung US | Bacterial Infection  n=67 | Viral Infection  n=76 | Atypical Infection  n=43 | *P value* |
| --- | --- | --- | --- | --- |
| Consolidation | 67 (100) | 65 (85.53) | 38 (88.37) | **B-V: 0.001**  V-A:0.66  **B-A: 0.004** |
| Size consolidation |  |  |  |  |
| <1.5 cm | 15 (22.39) | 41 (63.8) | 19 (50) | **B-V: < 0.001**  V-A: 0.19  **B-A: 0.004** |
| 1.5-4 cm | 37 (55.22) | 23 (35.38) | 17 (44.74) | **B-V: 0.022**  V-A: 0.347  B-A: 0.301 |
| >4cm | 15 (22.39) | 1 (1.54) | 2 (5.26) | **B-V: <0.001**  V-A: 0.306  **B-A:0.022** |
| Multiple consolidations | 5 (7.46) | 28(43.8) | 13 (34.21) | **B-V:< 0.001**  V-A: 0.375  **B-A:<0.001** |
| Bilateral consolidations | 6 (0.09) | 31(46.15) | 12 (31.58) | **B-V: <0.001**  V-A: 0.146  **B-A:0.004** |
| Air bronchogram | 52(77.61) | 25 (32.89) | 26 (60.47) | **B-V: <0.001**  **V-A:0.004**  B-A:0.053 |
| Position air bronchogram  Deep  Superficial | 28 (53.85)  24 (46.15) | 14 (56)  11 (44) | 6 (23.08)  20 (76.92) | B-V: 0.859  **V-A:0.016**  **B-A: 0.010** |
| Bronchogram  Dynamic  Static | 22 (44)  28 (56) | 9(37.50)  15 (62.50) | 18 (69.23)  8 (30.77) | B-V: 0.596  **V-A:0.025**  **B-A:0.037** |
| Fluid bronchogram | 22 (32.84) | 3 (3.95) | 4 (9.30) | **B-V: <0.001**  V-A:0.233  **B-A:0.003** |
| Pleural effusion | 33 (49.25) | 12 (15.79) | 7 (16.28) | **B-V: <0.001**  V-A:0.94  **B-A: <0.001** |
| Complicated effusion | 3 (10.34) | 0 | 2 (33.33) | B-V: 0.374  V-A: 0.192  BA: 0.143 |
| Vertical deep artifacts | 39 (58.21) | 67 (88.16) | 28 (88.37) | **B-V: <0.001**  V-A: 0.972  **B-A: 0.001** |
| Vertical deep artifacts  Diffuse  Peri-lesion | 17 (44.74)  21 (56.6) | 55 (78.57)  15 (21.43) | 26(70.27)  11(29.73) | **B-V: <0.001**  V-A:0.341  **B-A: 0.025** |
| Vertical deep artifacts  Confluent | 1 (2.63) | 4 (5.71) | 0 | B-V: 0.467  V-A:0.123  B-A:0.5 |

**Table 3.** Differences of the ultrasound features in community acquired pneumonia with three etiological causative agents.

| **B-V Infection** | **OR** | **z** | ***P*** | **95% (CI)** | |
| --- | --- | --- | --- | --- | --- |
| Gender (Female vs. Male) | 0.75 | -0.46 | 0.643 | 0.22 | 2.57 |
| Age (years) | 1.23 | 2.43 | 0.015 | 1.04 | 1.44 |
| Large sized consolidation | 13.62 | 2.08 | 0.038 | 1.16 | 159.88 |
| Multiple consolidation | 0.24 | -1.42 | 0.154 | 0.03 | 1.72 |
| Bilateral consolidation | 0.23 | -1.43 | 0.152 | 0.03 | 1.72 |
| Air bronchogram | 6.58 | 2.69 | 0.007 | 1.67 | 25.93 |
| Deep vertical artifacts | 0.27 | -1.88 | 0.060 | 0.07 | 1.06 |
| Pleural effusion | 1.48 | 0.61 | 0.543 | 0.42 | 5.16 |
| CRP>100 mg/L | 15.94 | 3-43 | 0.001 | 3.28 | 77.50 |
| Chest pain | 0.47 | -0.80 | 0.426 | 0.07 | 2.99 |
| Decreased air entry (auscultation) | 0.75 | -0.46 | 0.643 | 0.22 | 2.57 |
| Constant | 0.30 | -1.12 | 0.261 | 0.04 | 2.46 |

**Table 4**. Logistic regression model analyzing the diagnostic (clinical, laboratory and US) predictors for Bacterial Infection versus Viral Infection in community acquired pneumonia.

The model showed that the likelihood of the bacterial etiology increased significantly of more than 13 times for the large sized consolidation (p= 0.039; OR: 13.62; 95% CI: 1.16 to 159.88). The air bronchogram occurred more than 6 times in the bacterial community pneumonia. (p=0.007; OR: 6.58; 95% CI: .67 to 25.93). The CRP value of 100 mg/L was about 15 times as much associated with bacterial etiology (p=0.001; OR: 15.94; 95% CI: 3.28 to 77.50). In addition, the more age increased, the greater the likelihood of bacterial etiology (p= 0.015; OR: 1.23; 95% CI: 1.04 to 1.44).

OR: Odds Ratio; z: regression coefficient divided by its standard error; 96% CI: 95% confidence interval.

| **V-A** | **OR** | **Z** | ***p*** | **95% (CI)** | |
| --- | --- | --- | --- | --- | --- |
| Gender (Female vs. Male) | 0.41 | -1.35 | 0.176 | 0.11 | 1.49 |
| Age (years) | 0.93 | -1.00 | 0.319 | 0.80 | 1.07 |
| Air bronchogram | 0.14 | -2.84 | 0.004 | 0.04 | 0.55 |
| Cough | 0.33 | -1.25 | 0.210 | 0.06 | 1.87 |
| Respiratory distress | 8.35 | 1.80 | 0.073 | 0.82 | 84.56 |
| Crackles (at auscultation) | 0.58 | -0.80 | 0.426 | 0.15 | 2.23 |
| CRP <50mg/L | 4.15 | 1.74 | 0.082 | 0.84 | 20.66 |
| Constant | 13.27 | 2.47 | 0.014 | 1.70 | 103.56 |

**Table 5.** Logistic regression model analyzing the diagnostic (clinical laboratory and US) predictors

for Viral Infection versus Atypical Infection in community acquired pneumonia.

The model showed that the detection of air bronchogram at lung US reduced by 86% the odds of atypical CAP (p=0.004; OR: 0.14; 95% CI: 0.04 to 0.55).

OR: Odds Ratio; z: regression coefficient divided by its standard error; 96% CI: 95% confidence interval.

| **B-A** | **OR** | **z** | ***p*** | **95% (CI)** | |
| --- | --- | --- | --- | --- | --- |
| Gender (Female vs. Male) | 0.36 | -1.04 | 0.30 | 0.05 | 2.48 |
| Age (years) | 0.81 | -1.81 | 0.07 | 0.65 | 1.02 |
| Small sized consolidation | 0.82 | -0.18 | 0.86 | 0.09 | 7.65 |
| Air bronchogram | 0.41 | -0.72 | 0.47 | 0.04 | 4.57 |
| Pleural effusion | 12.70 | 1.88 | 0.06 | 0.89 | 180.29 |
| Bilateral consolidation | 0.26 | -0.59 | 0.56 | 0.00 | 24.26 |
| Multiple consolidation | 0.36 | -0.56 | 0.57 | 0.01 | 12.72 |
| Deep vertical artifacts | 1.00 |  |  |  |  |
| CRP < 50 mg/dl | 0.06 | -2.43 | 0.02 | 0.01 | 0.58 |
| Cough | 0.27 | -0.85 | 0.40 | 0.01 | 5.46 |
| Respiratory distress | 1.00 |  |  |  |  |
| Decreased air entry | 0.53 | -0.61 | 0.54 | 0.07 | 4.10 |
| Constant | 310.84 | 2.33 | 0.02 | 2.50 | 38716.21 |

**Table 6:** Logistic regression model analyzing the diagnostic (clinical, laboratory and US)

predictors for Bacterial Infection versus Atypical Infection in community acquired pneumonia.

The model showed that the value of CRP < 50 mg/dl reduced by 94% the odds of

bacterial etiology compared with atypical etiology (p=0.02; OR: 0.06; 95% CI: 0.01 to 0.58).

OR: Odds Ratio; z: regression coefficient divided by its standard error; 96% CI: 95%
